# A model for reconstructing trends and distribution in age at first sex from multiple household surveys with reporting biases

**DOI:** 10.1101/2021.12.14.21267770

**Authors:** Van Kính Nguyen, Jeffrey W Eaton

**Affiliations:** MRC Centre for Global Infectious Disease Analysis, School of Public Health, Imperial College London, London, United Kingdom

**Author notes:** Corresponding author: Van Kính Nguyen, MRC Centre for Global Infectious Disease Analysis, School of Public Health, Imperial College London, St. Mary’s Hospital Campus, Norfolk Place, London, W2 1PG.

**Keywords:** age at first sex, log-skew-logistic, age at report bias

## Abstract

Age at first sex (AFS) is a key indicator for monitoring sexual behaviour risk for HIV and sexually transmitted diseases. Reporting of AFS data, however, suffer social-desirability and recall biases which obscure AFS trends and inferences constructed from it. We illustrated examples of the biases using data from multiple nationally-representative Demographic and Health Surveys household surveys conducted between 1992 and 2019 in Ethiopia (4 surveys), Guinea (4 surveys), Senegal (8 surveys), and Zambia (8 surveys). Based on this, we proposed a time-to-event, interval censored model for the AFS that uses overlapping reports by the same birth cohort in successive surveys to adjust reporting biases. The three-parameter log-skew-logistic distribution described the asymmetric and nonmonotonic hazard exhibited by empirical AFS data. In cross-validation analysis, incorporating a term for AFS reporting bias as a function of age improved model predictions for the trend of AFS over birth cohorts. The interquartile range for the AFS was 16 years to 23 years for Ethiopian and Senegalese women and 15 years to 20 years for Guinean and Zambian men. Median AFS increased by around one to 1.5 years between the 1960 and 1989 birth cohorts for all four datasets. Younger male respondents tended to report a younger AFS while female respondents tended to report an older AFS than when asked in later surveys. Above age 30, both male and female respondents tended to report older AFS compared to when surveyed in their late twenties. Simulations validated that the model recovers the trend in AFS over birth cohorts in the presence of reporting biases. At least three surveys are needed to obtain reliable trend estimate for a 20-years trend. Mis-specified reference age at which reporting is assumed unbiased did not affect the trend estimate but resulted in biased estimates for the median AFS in the most recent birth cohorts.

## Introduction

Age at first sex (AFS) refers to the age when an individual begins sexual intercourse. This indicator has been routinely collected as part of national household surveys and used for monitoring efforts for prevention of HIV and sexual transmitted infections (STIs) as well as providing perspectives on social, gender equity, and demographic developments. Early AFS has been associated with higher rates of STIs (Pettifor *et al*., 2004; Kaestle *et al*., 2005) while delaying AFS decreases the risk of HIV infection (Hallett *et al*., 2007) and contributed to curtailing HIV epidemics (Halperin *et al*., 2004). Early AFS could also heighten the likelihood of engaging risky sexual behaviours that facilitate STIs (Kaplan *et al*., 2013a).

Data on AFS are typically collected via self-report in nationally-representative household surveys conducted roughly every five years to track health and population indicators, such as Demographic and Health Surveys (DHS) (ICF, 2021a) or Multiple Indicator Cluster Surveys (MICS)(UNICEF, 2021). Several challenges have been identified for measuring AFS. First, monitoring AFS has focused on summary statistics such as the median AFS indicator or the proportion sexually experienced by a particular age, for example by age 15 or age 18. These indicators do not characterise the full distribution, including tail behaviours, which inform transmission dynamics and identify sub-populations who are vulnerable to risk. Second, observations are right censored for respondents who have not had sex at the time of data collection. If not analysed appropriately, the observed median AFS is biased downwards for younger and more recent birth cohorts. A common strategy for calculating indicators is to exclude the youngest cohorts (for example respondents aged 15-19 years during the survey), but this discards information on more recent trends, which are likely of greatest public health interest.

Third, retrospectively reported AFS collected primarily through face-to-face interviews are susceptible to recall and social desirability biases. It can be difficult to accurately recall AFS that occurred many years ago and responses have been shown to be affected by desirable standards of society, culture, and health or political campaigns (Catania, 1999; Wellings *et al*., 2006; United Nations, 2012). Inconsistencies in AFS reports ranged 30%–56% between surveys in the same population (Eggleston, Leitch and Jackson, 2000; Wringe *et al*., 2009). There was also a tendency to report older AFS over time (Zaba *et al*., 2004a; Wringe *et al*., 2009) and as the cohort aged from teenager to adulthood, men reported higher AFS while women reported lower AFS (Mensch, Hewett and Erulkar, 2003; Zaba *et al*., 2004a).

In this paper, we elucidated reporting biases in empirical AFS data from multiple nationally-representative household surveys and developed and validated a survival model to estimate the distribution of AFS in a population over time, adjusting for previously described reporting biases.

## Materials and Methods

Our analysis consisted of four components. First, we conducted exploratory analysis of household survey data to elucidate the reporting biases described in previous literature and conceptualise bias adjustment models. Second, we empirically compared five alternative parametric survival distributions for representing the distribution of AFS. Third, we used cross-validation to compare model specifications for the survival distribution, trends by birth cohort, and bias adjustment components. Finally, we conducted simulations to assess situations where the model can reliably recover the trend in AFS and sensitivity to misspecification of model assumptions.

### Survey data

We obtained data on AFS from multiple Demographic and Health Surveys (DHS) conducted between 1992 and 2019 in Ethiopia (female respondents; 4 surveys), Guinea (male respondents, 4 surveys), Senegal (female respondents, 8 surveys), and Zambia (male respondents, 8 surveys). Individual-level survey responses were extracted on respondents’ age, year of birth, sex, and age at first sex. AFS was reported in integer ages, which we assumed to be the completed age at which sexual debut occurred. AFS was recorded as ‘never had sex’ for some respondents, forming right-censored observations at the age at interview. Respondents who reported an unknown AFS were dropped from analysis.

### Exploratory analysis

We explored reporting biases in AFS by comparing what the same birth cohort reported about their AFS in successive surveys. We calculated the proportion of respondents who had ever had sex before age 18 years among respondents aged 18 years and older stratified by respondents born 1970–1974, 1975–1979, 1980–1984, and 1985–1989. Estimates for the proportion and 95% CIs accounted for survey weights and clustered sampling design using ‘survey’ package in the statistical software R (Lumley, 2004).

### Parametric survival distributions

We considered five candidate parametric distributions to model the distribution of AFS for a birth cohort: the gamma distribution (two parameters), log-normal distribution (two parameters), log-logistic distribution (two parameters), generalised gamma distribution (three parameters), and log-skew-logistic distribution (three parameters). The log-skew-logistic distribution was derived from the skew-logistic distribution (also called generalized logistic distribution), defined by the density function

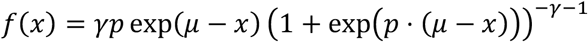

and the corresponding cumulative distribution

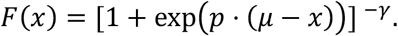

The parameters *μ, p* > 0, *γ* > 0 determine the scale, shape, and skewness of the distribution, respectively. The distribution reduces to a logistic distribution when *γ* = 1 (Appendix - Figure 2). The log-skew-logistic distribution was obtained by re-parameterising the scale parameter as *μ* = −log(*λ*) and performing a change of variable to *x* = log (*T*), the log AFS. The survival function is

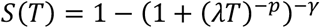

and density function is

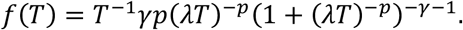

The hazard function for sexual debut at age *T* is

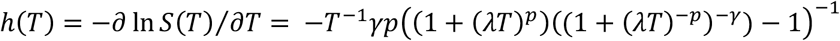

and the median AFS is 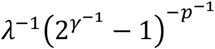.

To assess the suitability of the five distributions to model AFS distribution, we estimated the empirical smoothed hazard function and Kaplan-Meier estimator of the survival function for pooled survey datasets from each of the four countries. The smoothed hazard function was estimated using kernel-based methods with local bandwidth selection (Muller and Wang, 1994). Parameters were estimated by maximum likelihood for each of the five parametric survival models. The resulting hazard and survival function were compared visually to the corresponding non-parametric estimates.

### Survival model for time to AFS

AFS data were recorded as two variables: an indicator of whether an individual had ever had sex and an integer variable indicates at what age. Let *T*_*i*,_ *i* = 1, …, *n* the AFS of individual *i*, and *d*_*i*_ an indicator for whether the respondent reported ever having had sex such that *d*_*i*_ = 1 indicated a reported AFS; those who answered never had a sexual intercourse were assumed right censored with *d*_*i*_ = 0. For censored cases, *T*_*i*_ was specified as the age at the time of interview.

We assumed that AFS was reported as the completed years at last birthday when first intercourse occurred. Thus, reported AFS was not interpreted as exact ages, but as interval censored observations surrounding the true AFS 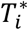, that is, 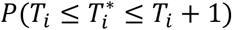. The probability of AFS observations can be described by an interval censored survival model with the likelihood as

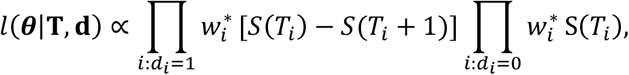

in which *S*(·) is the survival function of a selected distribution for AFS. The weights 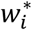 reflect unequal sampling weights determined by the survey sampling design. For each survey *k*, the raw weight *w*_*ik*_ for observation *i* was scaled as 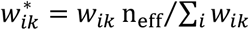 where 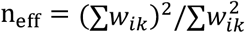 is the Kish’s effective sample size (Leslie Kish, 1995), which accounted for the loss of sampling efficiency due to heterogeneous sampling weights.

### Model specification for time trends and reporting biases

The five distributions considered can be characterized mainly with a scale parameter which controls how spread out or concentrated the distribution. Depending on the distribution, other parameters including shape and skewness give finer control of the distribution’s shape and behaviours of the tails. The effects of covariates, including age and birth cohort, was modelled on the distribution’s scale parameter. We estimated

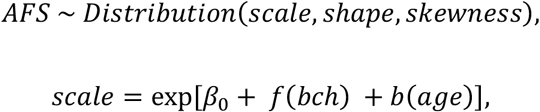

where *β*_0_ the intercept, *f*(*bch*) denotes effect of the birth cohort and *b*(*age*) the bias to the scale by the age at report. We considered both birth cohort and age at report effects can be linear or nonlinear. Nonlinear effects were specified with a first-order random walk (RW1) structure. The standard normal was used as prior for the intercept and coefficients of birth cohort and age at report in linear form. The prior distribution for the random walk’s precision parameters was specified using the penalized-complexity prior (Simpson *et al*., 2017). Sum-to-zero constraints were imposed on each of the random walk component in consideration, that is, Σ_*age*_ *b*(*age*) = Σ_*yop*_ *f*(*bch*) = 0. The reference age for the reporting bias term *b*(*age*), at which reporting was assumed to be accurate, was age 23.

### Model implementation and comparison

The model was implemented with Template Model Builder (Simpson et al., 2017). Posterior distributions for model parameters were estimated with empirical Bayesian framework using Laplace approximation for the marginal likelihood. Ten-folds cross-validation was conducted to compare distributions and model structures in real survey datasets. The data was randomly divided to ten subsets; the model was fitted with one of the subsets omitted each time and the resulted model was used to compute the out-of-sample expected log-likelihood of the omitted subset. The expected log pointwise predictive density (ELPD) defined as (Vehtari, Gelman and Gabry, 2017)

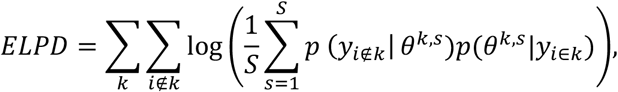

was reported where *S* = 1000 is the number of draws from the parameter’s posterior distribution estimated from all partitions excluding partition *k, k* = 1,…, 10. The larger the ELPD the better the model in consideration. Code is available at https://github.com/kklot/dbmethod.

### Simulation study

To characterise the model’s ability to capture true AFS, simulated datasets were generated reflecting alternative specifications about biases, true trends, and number of available surveys. We challenged the model to recover AFS of birth cohorts spanning from 1985 to 2005 with 2, 3, 4, or 5 surveys conducted between 2001 and 2020. The steps to simulate the survey data were: (i) generate a pooled population of 1 million with year of birth assigned uniformly between 1940–2005, (ii) for each birth cohort, assign a true AFS distribution using the log-skew-logistic distribution; the parameters were chosen such that the median AFS increased from 15 to 19, decreased from 19 to 15, or constant at 17 from 1940 to 2005, (iii) pick *k* = 2, . .,5 surveys conducted five years apart, started from 2020 and went backward in time. This ensures the target birth cohorts, 1985–2005, was fully covered in one survey and partially in the others with progressive lesser information about recent birth cohorts. In each survey, 1000 adults aged 15–49 at the time of survey were sampled with probability of selection proportionate to a DHS dataset’s age frequency (Appendix - Figure 1), (iv) add a reporting bias to the true median AFS in step (ii); the bias pattern was a function of age of the respondent at the time of the survey and varied depending on the scenarios under assessment. The reported AFS for each individual was sampled from the biased parameters.

In addition to scenarios without reporting biases, two bias patterns were considered: a bias towards misreporting an earlier AFS by young adult respondents, as might be expected by young men reporting earlier sexual experience (Figure 1A.1) and bias of younger adults reporting an older AFS, as expected if young women tended to misreport abstaining sex until marriage (Figure 1B.1). Figure 1 shows example of changes in median AFS that would be reported by each cohort in four surveys 5-years apart under each bias pattern under a scenario with no true change in AFS over time.

**Figure 1.**
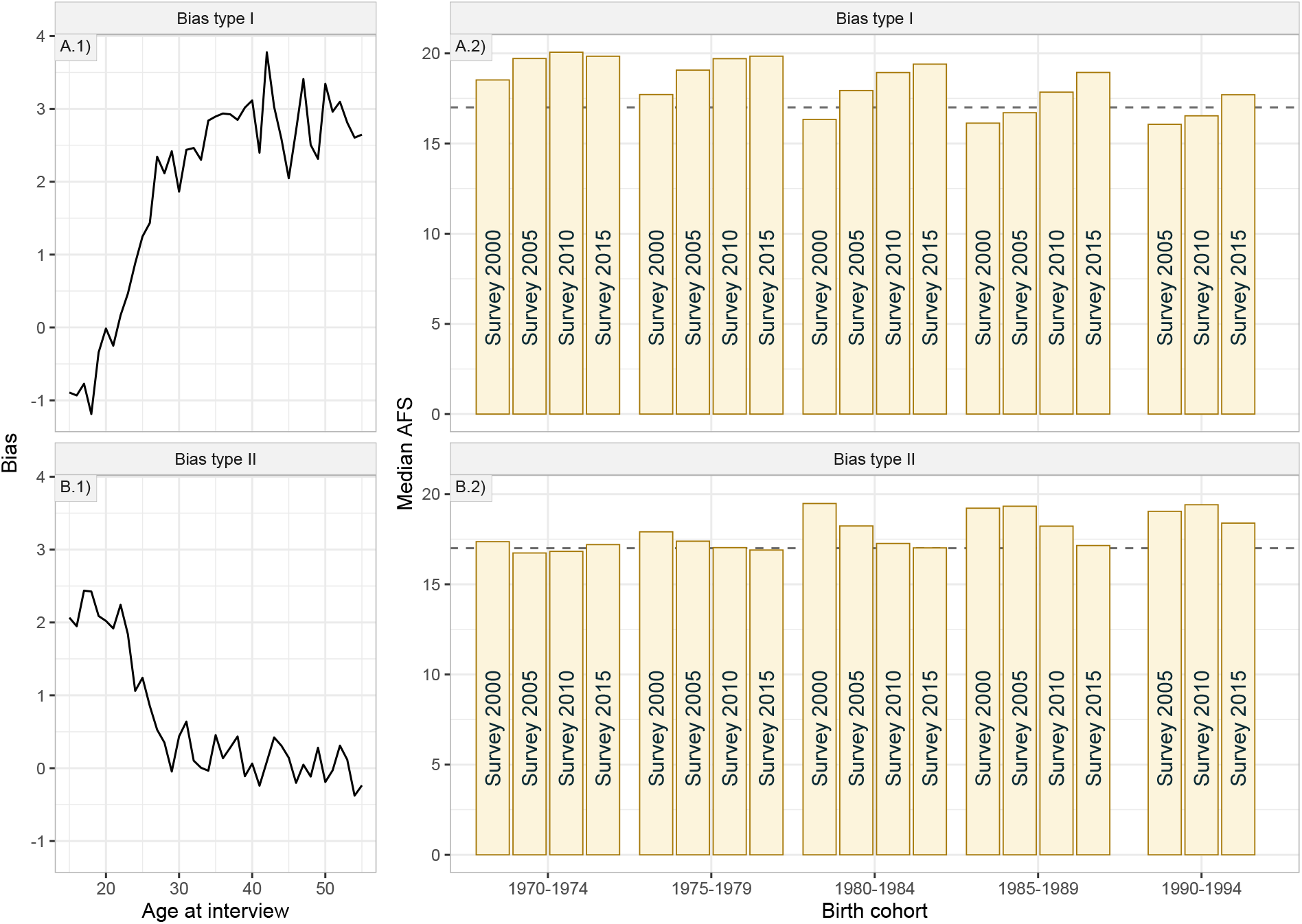
Simulated survey datasets of AFS (A.2, B.2) from two types of potential biases in reporting age at first sex (AFS) assuming different bias function of age at interview: A.1) exaggerating ever had sex by reporting younger AFS in younger ages, no longer do so at certain age (23-year-old), and starting to recall an older AFS afterward, B.1) hiding ever had sex by reporting older AFS in younger ages but no longer do so in older ages. The horizontal dashed line is the true AFS used in the simulations.

Each survey scenario was simulated 1000 times; each time, we estimated the model’s parameters and reported the average difference from the true median AFS based on 1000 posterior samples. To measure if the simulated trend was captured, the absolute difference between the estimated median AFS for the 1985 and 2005 birth cohort was reported. We also reported the error in the estimated median AFS of the most recent birth cohort for its pertinent in AFS communication.

### Ethics statement

The study involved secondary analysis of anonymised publicly available data from the Demographic and Health Survey programme following project approval (https://dhsprogram.com/data/available-datasets.cfm). Primary survey protocols were reviewed and approved by the Institutional Review Board of ICF International and the relevant national ethics committee in each country (https://dhsprogram.com/methodology/Protecting-the-Privacy-of-DHS-Survey-Respondents.cfm).

## Results

### Within-cohort inconsistency in reported AFS

Figure 2 shows the crude percentage who reported ever having had sex before age 18 years by birth cohort in successive surveys among respondents aged 18 and above. In all four countries, the percentage who reported sexual debut before age 18 varied by up to 10%–20% for the same cohort in successive surveys, indicating inconsistent reporting within the cohort. For men in both Guinea and Zambia, the proportion who reported sex before 18 reduced steadily as the cohorts aged, indicating that either younger men systematically over-reported their sexual activity or older men underreported. In Gambia the reduction was over 30% from the first survey in 1999 to the most recent in 2018. For women in Ethiopia, the percentage within each cohort reporting sex before age 18 consistently increased in successive surveys, contrasting the pattern among men. The largest increase was as women aged from their early 20s to early 30s, with smaller changes thereafter. This pattern was consistent with the youngest women under-reporting sexual activity. In Senegal, there was some evidence that the proportion reporting AFS before 18 increased as cohorts aged through their 20s, but changes were smaller and less consistent than for women in Ethiopia.

**Figure 2.**
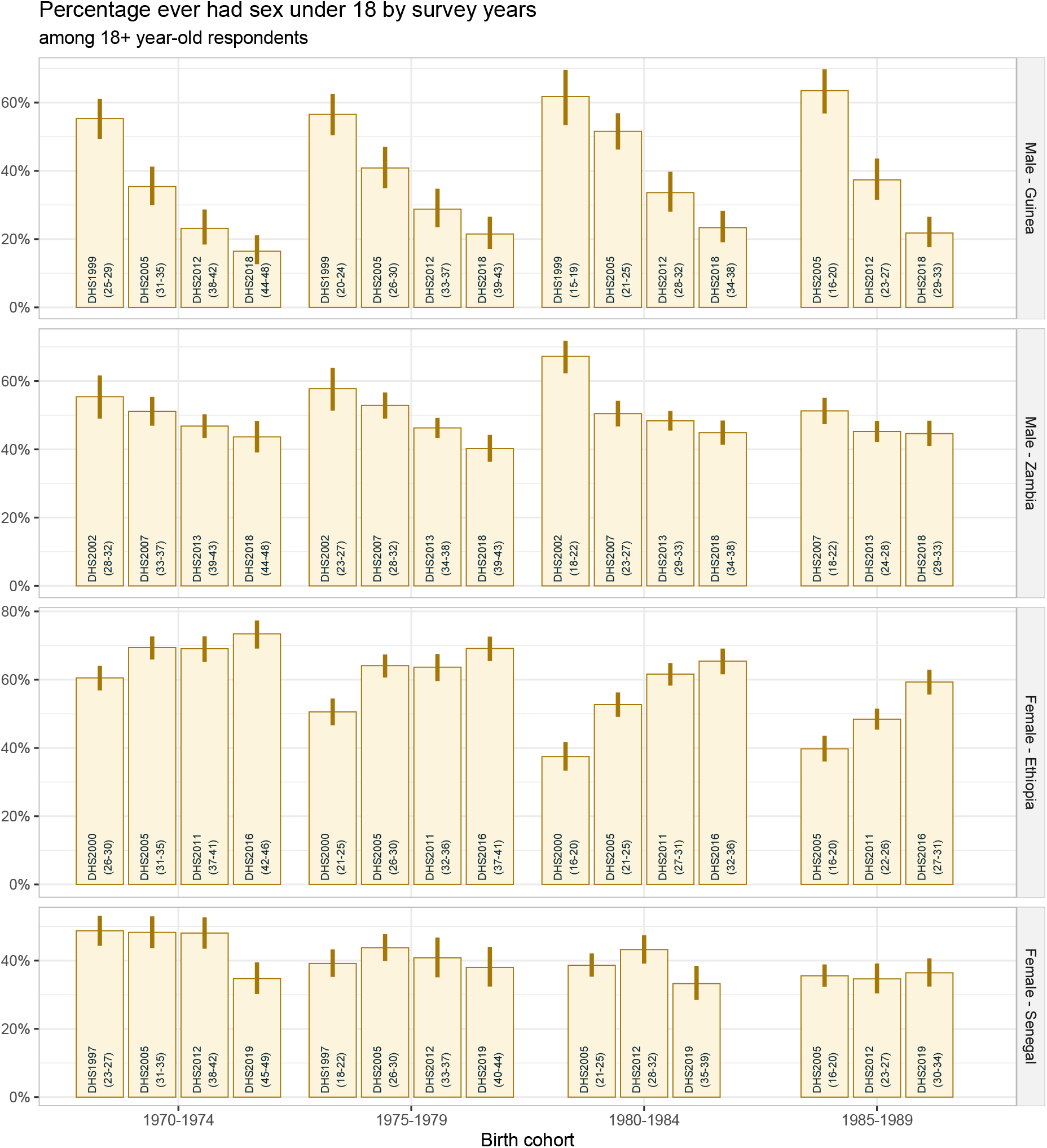
Within-cohort inconsistency in reported AFS over survey years across countries. The column’s label indicates the survey year and age at report in the bracket. Bars present point estimate and lines range are 95% confidence interval. The presented Senegal’s surveys are selected equally distant from each other started from the earliest survey.

### Parametric distribution for AFS

The empirical hazard rate of sexual debut initially increased slowly from age 10 to 13, then more rapidly to age 18 to 20, and stabilised between 0.2 and 0.3 per year (Figure 3–dashed black line). Above age 25, the empirical hazard declined slightly for women, but by this age above 85% to 90% had initiated sexual activity. The Gompertz and Weibull distributions were not considered here because both imply an exponentially increasing hazard function, which was not suitable for these data. The best-fitting gamma, log-normal, and generalized gamma distribution did not permit the flattening and decline of sexual debut hazard above age 20 (Figure 3). The log-logistic distribution and log skew-logistic were more consistent with the empirical hazard for AFS, though did not fully capture the decline. We limited further assessments to the log-logistic and the log-skew-logistic distribution.

**Figure 3.**
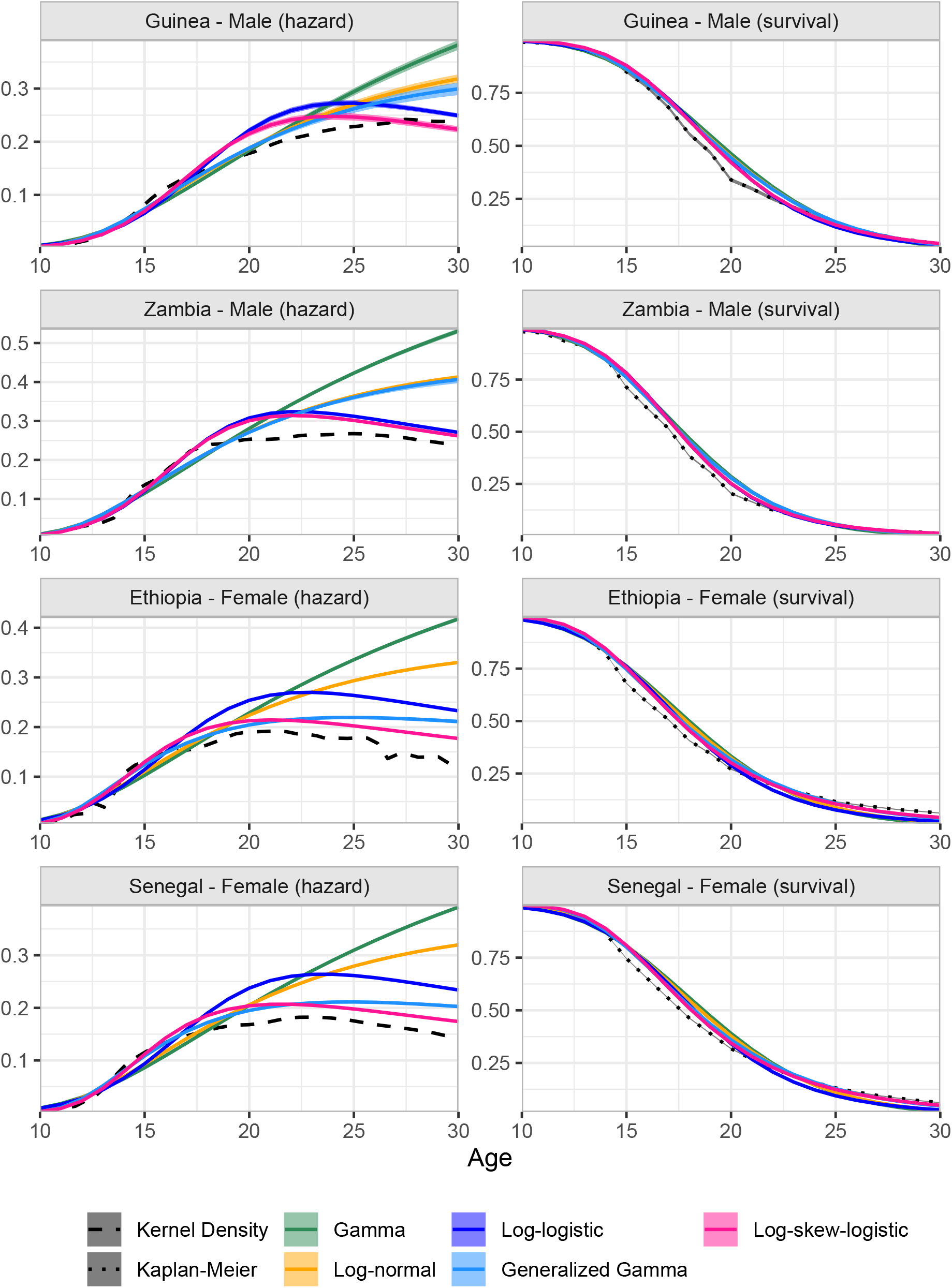
Fitted non-parametric (kernel density) and intercept-only model of parametric hazard functions of the common survival distributions to AFS data. The corresponding estimates of the survival function are shown on the right panel.

**Figure 4.**
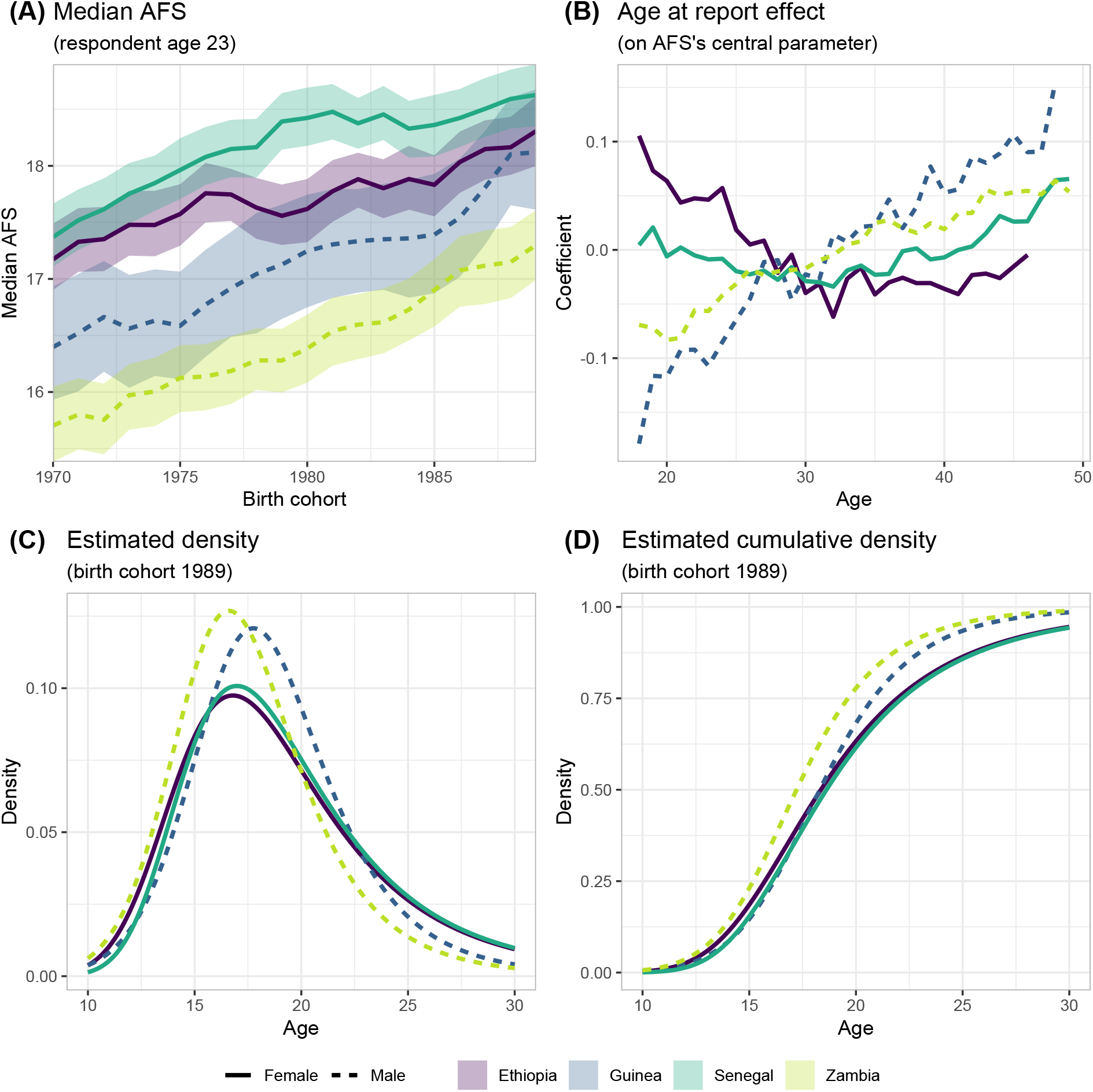
Estimated median AFS, log-skew-logistic density distribution, and age at report effect from the best model based on ELPD (Table 1). Age at report effect is fixed within birth cohort and directly change the scale parameter on the log-skew-logistic distribution.

### Comparison of modelled survival distributions and structures

In 10-folds cross-validation, the best model, measured by the model with the highest ELPD, was Model 5 using the log-skew-logistic distribution (Table 1). Model 5 included a smoothed term (RW1) for both birth cohort trend in AFS and a smooth RW1 term for the bias in reported AFS by age at report. The log-skew-logistic distribution provided better predictions than the log-logistic distribution for all four datasets and all model specifications. The difference was largest for the Senegal and Ethiopia female datasets, for which including skewness allowed lower sexual debut rates in very young ages followed by a more rapid increase to the peak of the distribution. Introducing the age at report bias term effected a large increase in the predictive value, with the nonlinear RW1 form preferred over a linear age at report effect.

**Table 1.** 10-folds cross-validation of the log-logistic and its skewness extension version. The out-of-sample expected log pointwise predictive density (ELPD) is computed for 1000 draws from the parameters posterior distributions. The values presented are the difference to the best model which is showed as zero EPLD in each country and weighting version. The plus sign preceeding the term denotes a model with intercept and the term, two pluses sign (++) denotes model inlcude intercept, RW1 effect of birth cohort (BCH), and the effect of age at report (AAR) whether as fixed effect of as RW1 random effect.

|  | Model | Log-logistic | Log-skew-logistic |
| --- | --- | --- | --- |
| Ethiopia<br>Female<br>(n=59710) | M1: Intercept | -4352.4 (607.7) | -3187.8 (601.84) |
|  | M2: + Birth cohort linear | -1808.8 (636.0) | -793.60 (622.47) |
|  | M3: + Birth cohort RW1 | -1658.6 (603.0) | -630.22 (623.47) |
|  | M4: + Birth cohort RW1 + age at report linear | -1235.6 (625.0) | -273.69 (614.38) |
|  | M5: + Birth cohort RW1 + age at report RW1 | -931.0 (637.9) | 0 (0) |
| Guinea Male<br>(n = 12936) | M1: Intercept | -721.46 (197.13) | -674.5 (199.67) |
|  | M2: + Birth cohort linear | -539.84 (194.07) | -505.13 (194.91) |
|  | M3: + Birth cohort RW1 | -451.07 (196.39) | -416.16 (196.95) |
|  | M4: + Birth cohort RW1 + age at report linear | -28.39 (193.14) | -5.64 (193.14) |
|  | M5: + Birth cohort RW1 + age at report RW1 | -21.77 (193.63) | 0 (0) |
| Senegal<br>Female<br>(n = 110657) | M1: Intercept | -5120.92 (581.69) | -2073.61 (589.81) |
|  | M2: + Birth cohort linear | -3196.61 (598.73) | -418.39 (605.22) |
|  | M3: + Birth cohort RW1 | -3033.72 (608.66) | -279.88 (596.65) |
|  | M4: + Birth cohort RW1 + age at report linear | -3036.15 (609.98) | -277.38 (600.58) |
|  | M5: + Birth cohort RW1 + age at report RW1 | -2713.72 (610.29) | 0 (0) |
| Zambia Male<br>(n = 37001) | M1: Intercept | -441.97 (319.10) | -412.21 (319.71) |
|  | M2: + Birth cohort linear | -424.85 (317.90) | -393.88 (317.57) |
|  | M3: + Birth cohort RW1 | -404.01 (318.15) | -375.63 (318.7) |
|  | M4: + Birth cohort RW1 + age at report linear | -46.84 (316.18) | -1.11 (316.96) |
|  | M5: + Birth cohort RW1 + age at report RW1 | -44.82 (315.45) | 0 (0) |

### Estimates of AFS trend and bias pattern

Figure 3 shows results of the best fitted model from 10-folds cross-validation which used the log-skew-logistic distribution for AFS, and RW1 models for both the trend by birth cohort and age at report bias terms for the four DHS datasets. Results were centred at reporting age of 23 years, which is around the median age at first marriage for men (23 to 24 years) and above age at first marriage for women (16 to 20 years) in these countries (ICF, 2021b).

Among the four datasets analysed, the estimated median AFS was highest for Senegalese women and lowest for Zambian men and increased by around one to 1.5 years between the 1960 and 1989 birth cohorts (Figure 3A). For the 1989 birth cohort, the rate of sexual debut peaked around age 16 or 17 (Figure 3C). The interquartile range for the AFS was 16 years to 23 years for Ethiopian and Senegalese women and 15 years to 20 years for Guinean and Zambian men (Figure 3D).

The pattern of age at report bias on the reported AFS varied by sex for young adults (Figure 3B). Consistent with the descriptive results in Figure 2, at younger ages, male respondents tended to report a younger AFS while female respondents tended to report an older AFS than when asked in later surveys. Above age 30, both male and female respondents tended to report older AFS compared to when surveyed in their late twenties. Figure 5 presents an example of the effect of adjusting for age at report on model fit to the reported percentage ever had sex by age 18 across the four surveys conducted in Guinea between 1999 and 2018; the predicted values of a model with and without considering age at report, showing that the shifts observed in the AFS data across the four surveys can be explained by the difference in age at report.

**Figure 5.**
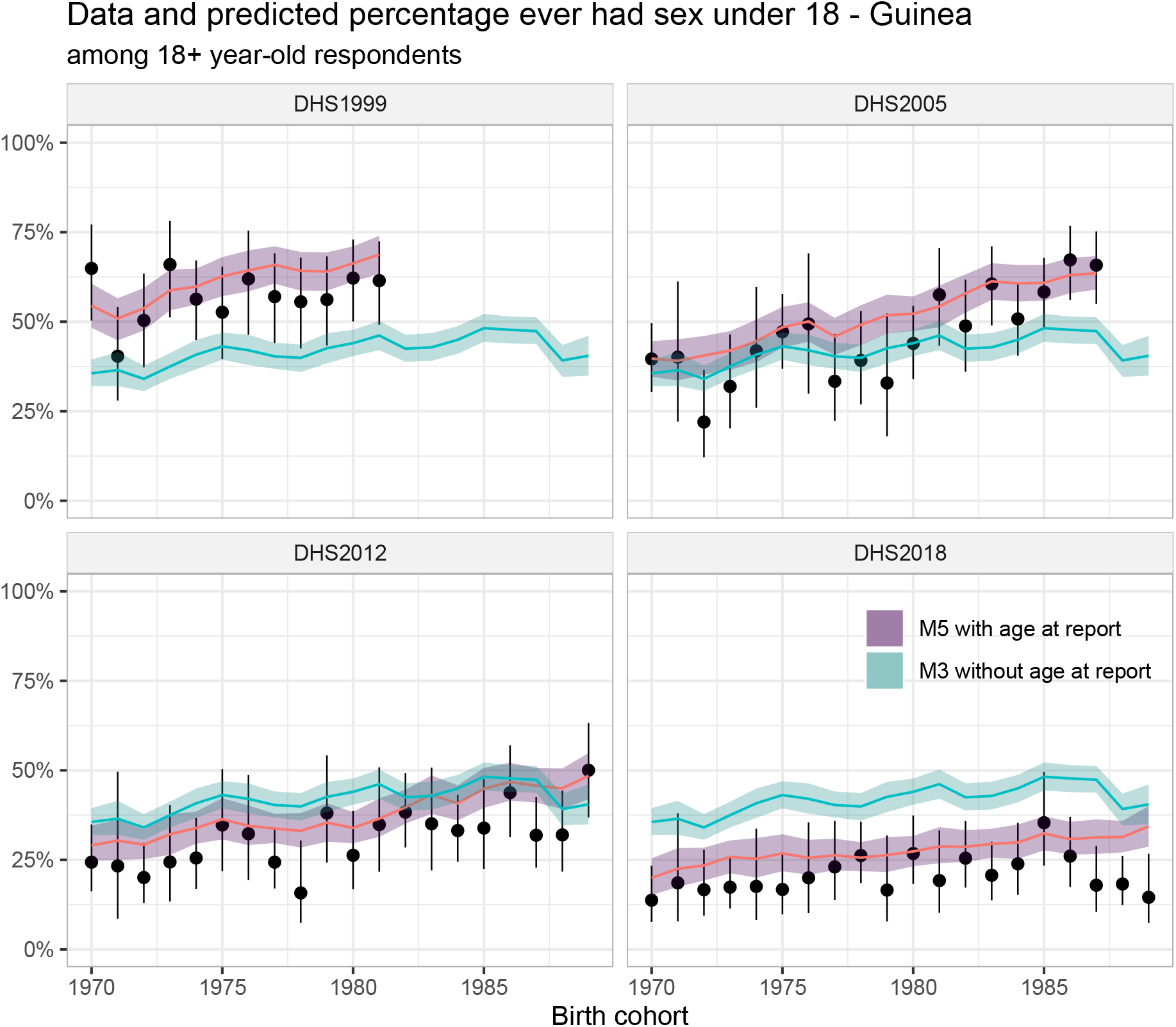
Illustration of predicted values of the model with and without taking account the age at report effect among male respondents in four DHS surveys in Guinea. Model M3 includes a RW1 term for the birth cohort but with the age at report effect ignored, model M5 is model M3 plus a RW1 model of the age at report.

### Sensitivity of model to survey settings and biases

Simulation study results for the change in the median AFS between the 1985 and 2005 birth cohorts are shown in Figure 6 for scenarios varying the number of surveys, trends, and bias patterns**Error! Reference source not found**.. The model identified the existence of a trend across the scenarios including when the data were biased or the age without bias was mis-specified. Average percentage of simulations correctly identified change in median AFS of more than one-half year ranged 67%–99% when three or more surveys were available (**Error! Reference source not found**.). Using only the two most recent surveys was prone to have incorrect estimates of trend. For example, when there was reporting bias and no trend, estimates from two sets of surveys suggested a trend of less than a year (Figure 6) and only correctly identified the decrease trend 34% of the times when data had the bias of young respondents reporting older AFS. With 4 or 5 surveys (providing more information for the older birth cohorts) the percentage increased to 85%–99%. The model was sensitive in detecting trend but underestimated the true magnitude of change when fewer surveys were available, but little was gained from increasing the number of surveys from 4 to 5. Figure 6 shows that mis-specified the reference age-group at which reporting was assumed to be unbiased did not affect the trend estimate, but it gave incorrect estimates for the true median AFS when there were biases (Appendix - Figure 4). The figure also shows that the magnitude and age pattern of biases were recovered correctly.

**Figure 6.**
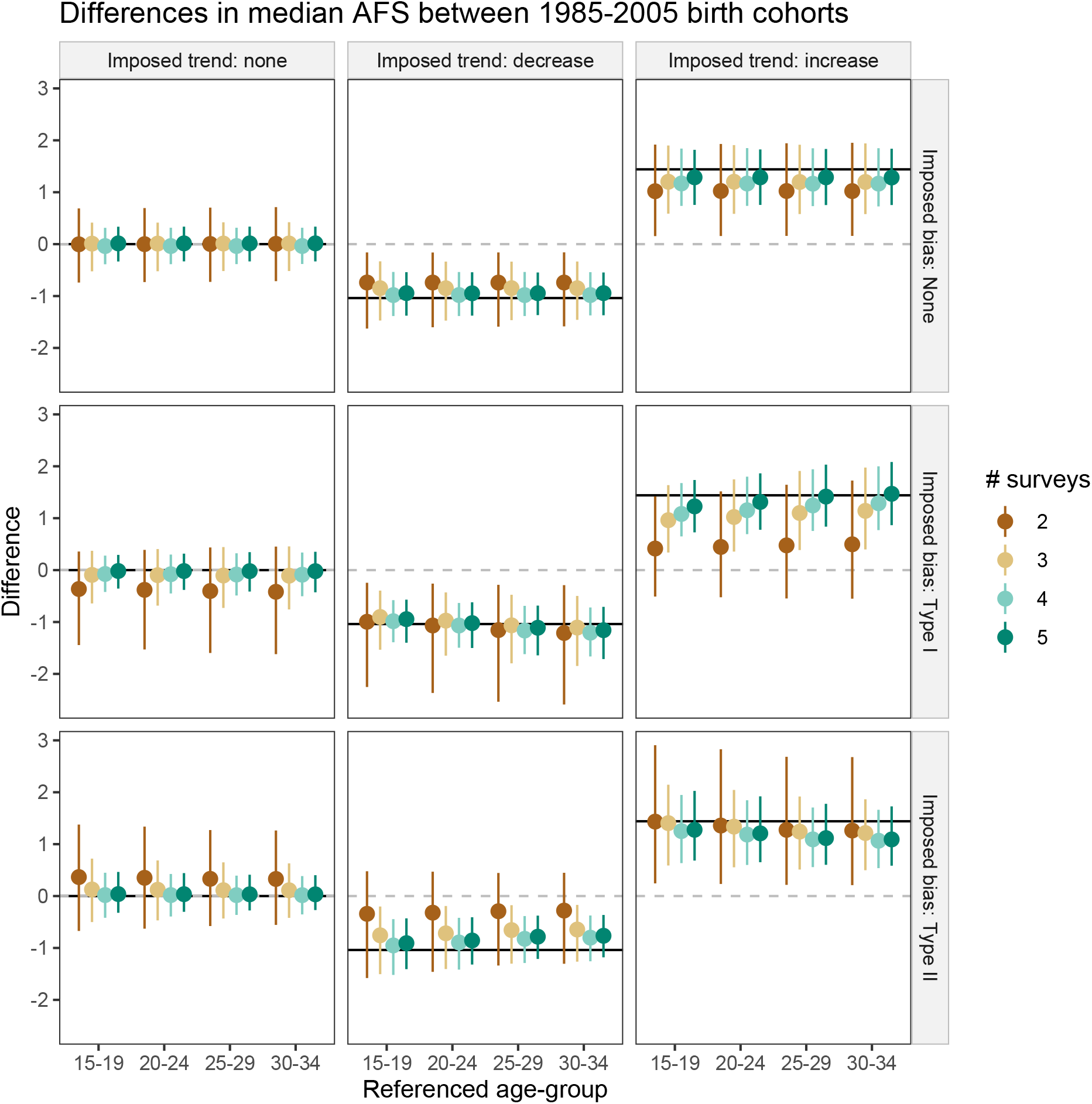
Estimated level of changes in the median AFS between the 2005 and 1985 birth cohort. The solid lines are the true difference imposed in the simulation. The type of biases are showed in Figure 1). The age’s effect reference group in the x-axis illustrates the effect on the difference when the average age coefficients of ages in the group was used in generating the AFS distribution.

## Discussion

Across many settings, changes in age at first sex (AFS) have been key determinants of changes in adolescent pregnancy and sexually transmitted disease, especially HIV/AIDS. However, measuring changes of this indicator over time is challenging due to biases arising from societal, religious, and cultural norms on sexual behaviours. In this study, we elucidated cases of potential biases in population surveys data of AFS and whether we could realize them in the analyses.

In the example datasets, large within-cohort inconsistencies across surveys four to six years apart implied different patterns of sexual behaviours from one survey to the next. Differences were less pronounced between surveys conducted annually in Senegal. Analyses of survey data conducted every two years apart also found small to no within-cohort differences (Wringe *et al*., 2009). In these cases of overlapping of respondents between surveys, repeatedly asked about AFS in a year interval might prompt respondents to provide similar answer. The varying within-cohort inconsistency across countries and sexes suggested that bias pattern is sex- and area-specific. This might be attributed to differences in social and cultural norms regarding desired sexual behaviours. Models attempting to capture this effect will need to permit sex and country-level variations of the within-cohort inconsistency.

At each survey year, age eligibility restricts the coverage of the birth cohorts included in the sample. For example, a survey conducted in year 2020 will sample the 2000-2005 birth cohorts as they progress through sexual debut. Statistics on age at first sex indicators must be computed accounting for censoring of the AFS data, for example, only including data from cohorts that have largely completed sexual debut when computing indicators on the percentage of ever had sex, obtaining median estimate from a Kaplan-Meier estimator, or modelling the data in a survival framework.

Previous studies have applied survival modelling framework in estimating AFS using the Gamma and generalized Gamma distribution (Zaba *et al*., 2004a; Fagbamigbe and Idemudia, 2017) but had not considered the asymmetric hazard of sexual debut distribution. In all our example datasets, the empirical estimate of the sexual debut hazard declined with age above age 20, which ruled out many distributions that did not permit this pattern. Cross-validation results showed that the asymmetric log-skew-logistic distribution reproduced empirical AFS data better than commonly used distributions.

Literature has identified many other factors that affect the AFS such as place of residence (Janis, Ahrens and Ziller, 2019), education (Hargreaves *et al*., 2012), ethnicity (Kaplan *et al*., 2013b), and religious belief (Rostosky *et al*., 2004). While these factors could explain differences between surveys, they represent subgroup differences compared to the systematic inconsistency arose from inherent age differences between surveys rounds (Eggleston, Leitch and Jackson, 2000; Zaba *et al*., 2004b). Leveraging these age differences to model the biases proved to be highly beneficial in improving model’s predictive ability as demonstrated in our cross-validation result. Furthermore, the estimated bias pattern from the examples data formed coherent mechanisms as has been theorized in literature.

Simulation results validated that the bias pattern on age can be recovered across the bias types and data trends (Appendix - Figure 4), and by using three or more surveys, the trend recovered was also more likely correct (Figure 6). As countries are progressing in monitoring and conducting more national surveys, there will be few countries with only one or two surveys in the future. As of 2021, however, there are still many countries in Sub Saharan Africa with sparse data on the AFS, e.g., Angola, Central African Republic, Comoros, Eritria, Guinea-Bissau, Sudan, and South Sudan, among others. In these cases, simultaneously model the effect of age at report using spatial model could help to guide the estimate in countries with few surveys available. The effect nonetheless needs to be flexible enough to permits country-specific difference as observed in this paper.

Simulation also showed that misspecification of the reference age at which AFS reporting is unbiased produced correct estimate of the overall trend but biased estimates of the AFS. It is practically difficult to ascertain the correct age of reference, which might also vary between countries. If there exist only social-desirability and recall biases, selecting an age when most of the respondents are expected to have had sexual debut, such as after median age at marriage, and not be too far from the median AFS would reduce potential bias introduced by misspecifying. Furthermore, ages that are known to have other type of biases such as digit preferences (Camarda, Eilers and Gampe, 2008) would be considered to be excluded.

The are several limitations in this study. First, as we limited the assessments to the age at report bias mechanism informed by previous empirical research (Zaba *et al*., 2004b), there are other types of biases which were not considered, for example temporal trends in reporting biases across all age groups. Second, we considered several commonly used distributions in survival analyses but might unknowingly leave out distributions with similar form of hazard or consider semi- or non-parametric hazard models. A parsimonious parametric hazard model is convenient for regression analyses or as inputs more complex models such as demographic or infectious disease models. Third, while the AFS distribution’s location parameter was allowed to vary over time, the skewness and shape parameter were fixed to borrow information from birth cohorts with a more complete AFS profile for the censored birth cohorts.

In conclusion, the distribution of age at first sex is asymmetric, and its hazard is nonmonotonic which can be described with the log-skew-logistic distribution. Estimating AFS trends and magnitude should account for potential biases according to the studied context. Age at report bias could mislead the current estimates of the AFS, however modelling data from several repeated surveys can help to identify the true underlying trend.

## Data Availability

All data produced are available online at https://dhsprogram.com/data/Dataset-Types.cfm

## Acknowledgements

VKN and JWE conceived the work. VKN and JWE designed the work. VKN implemented the models. Both authors critically reviewed model results throughout the model development process. VKN wrote the first draft of the manuscript. Both authors critically edited the manuscript for intellectual content.

This research was supported the Bill and Melinda Gates Foundation (OPP1190661), National Institute of Allergy and Infectious Disease of the National Institutes of Health under award numbers R01AI136664 and R01AI152721, and the MRC Centre for Global Infectious Disease Analysis (reference MR/R015600/1), jointly funded by the UK Medical Research Council (MRC) and the UK Foreign, Commonwealth & Development Office (FCDO), under the MRC/FCDO Concordat agreement and is also part of the EDCTP2 programme supported by the European Union.

## Appendix

**Appendix - Figure 1.**
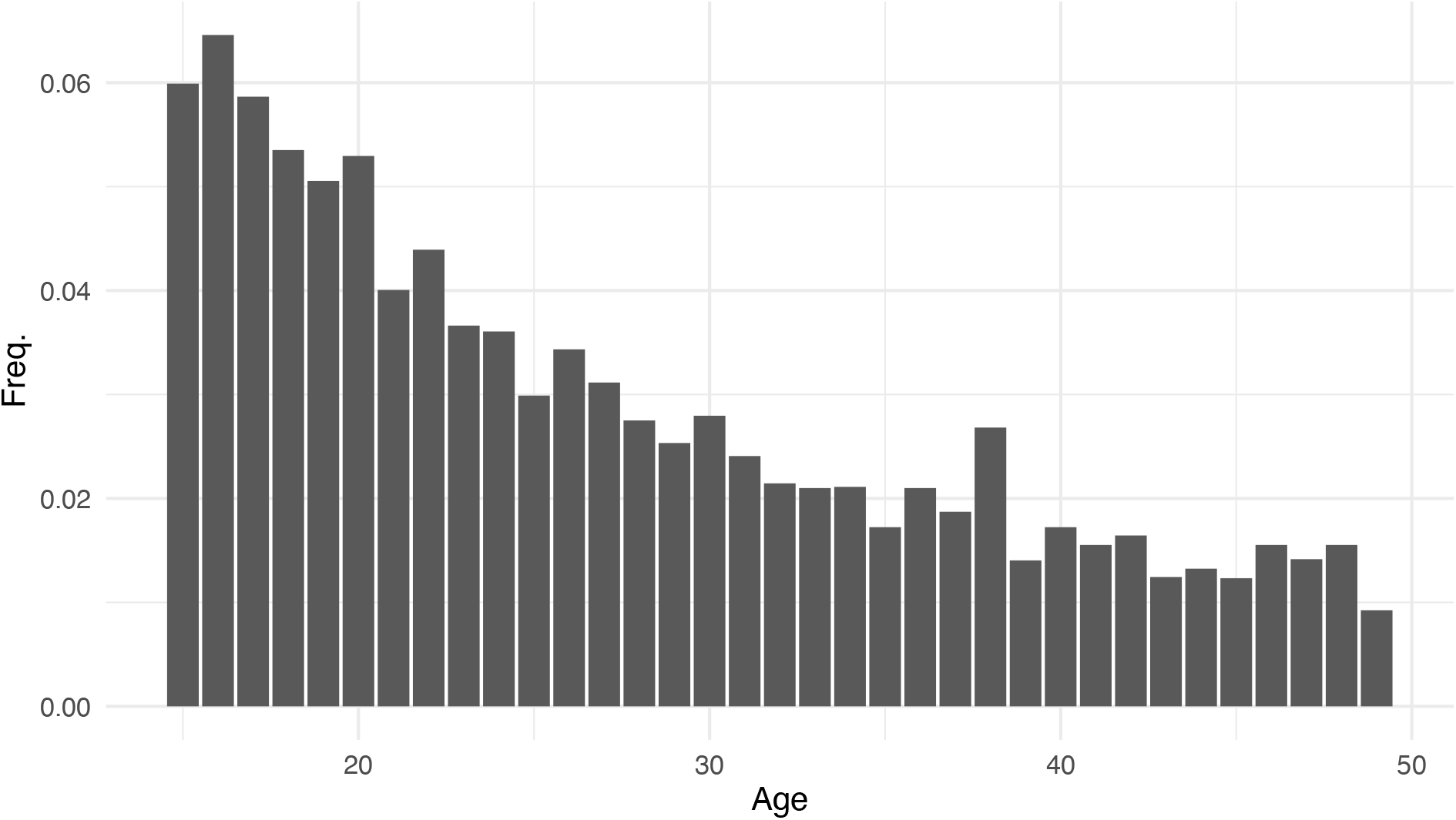
Age distribution of Eswatini’s DHS survey used in generating simulated survey datasets.

**Appendix - Figure 2.**
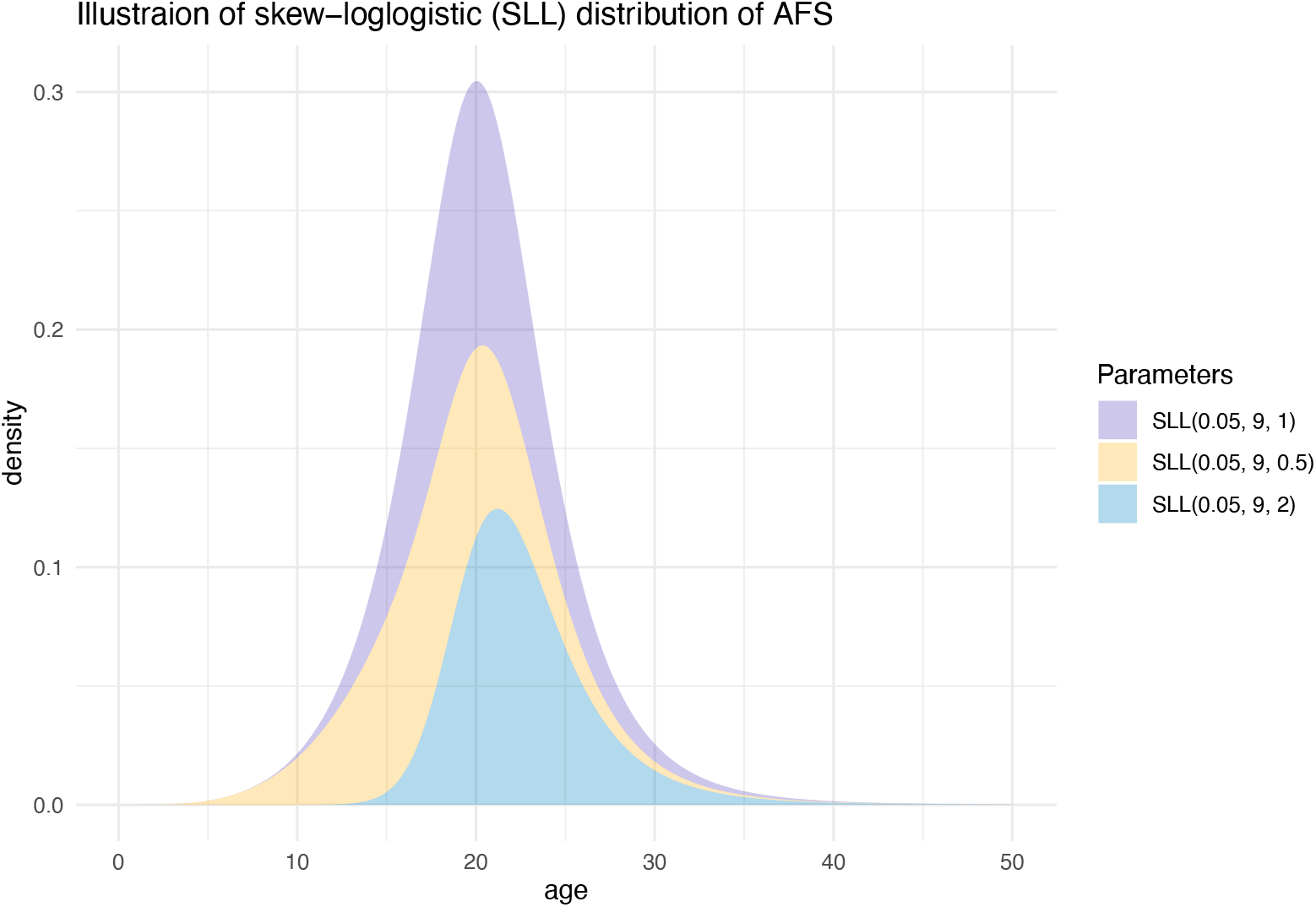
Illustration of log skew-logistic distribution with different levels of skewness in the data.

**Appendix - Figure 3.**
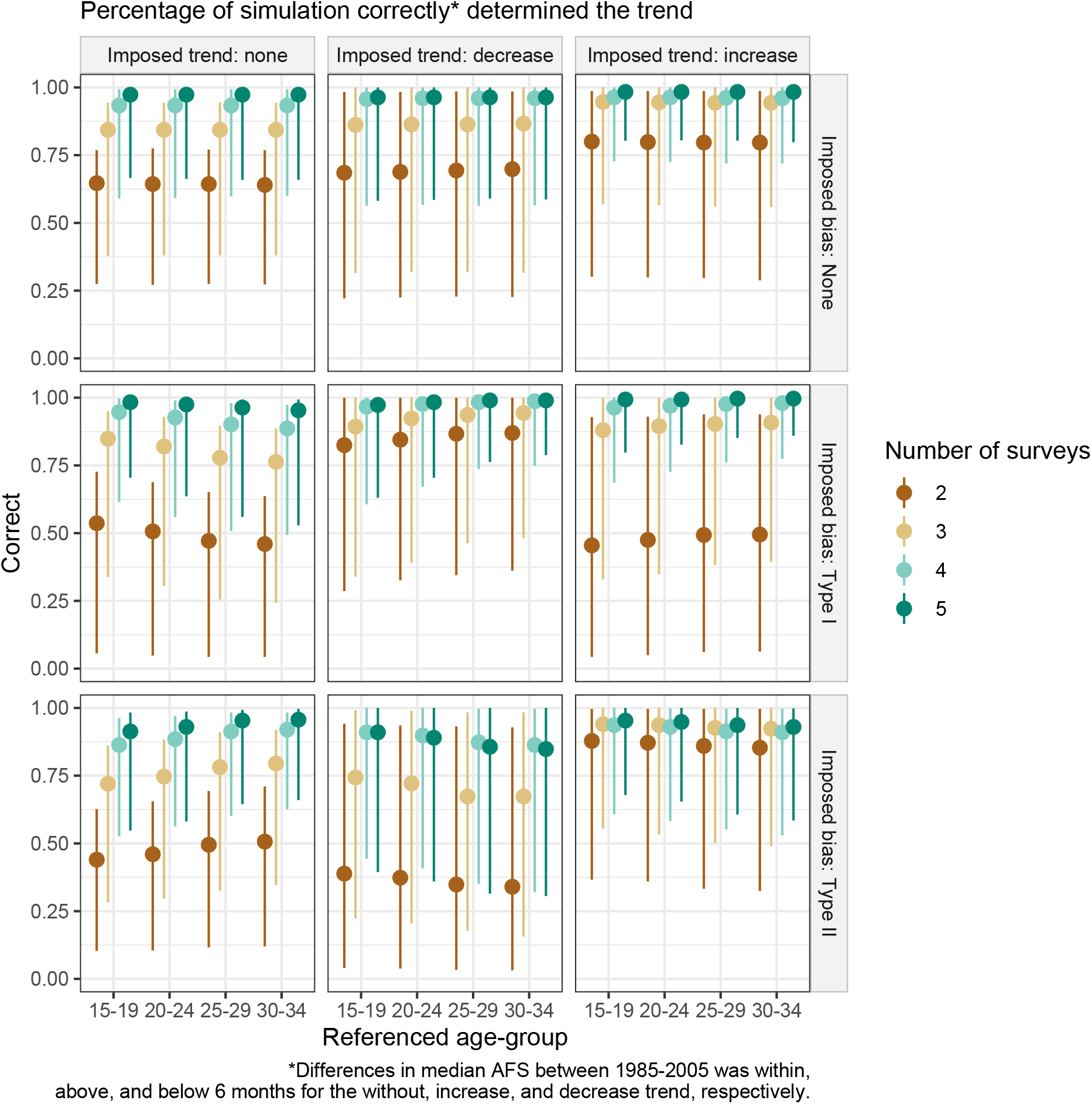
Percentage of simulation correctly determined the trend imposed in the simulated dataset. The trend was determined correctly when the model detected a change of more than 6 months in median AFS when there was a trend imposed; changes in median AFS of less than 6 months in either of the directions were considered as no trend in this figure.

**Appendix - Figure 4.**
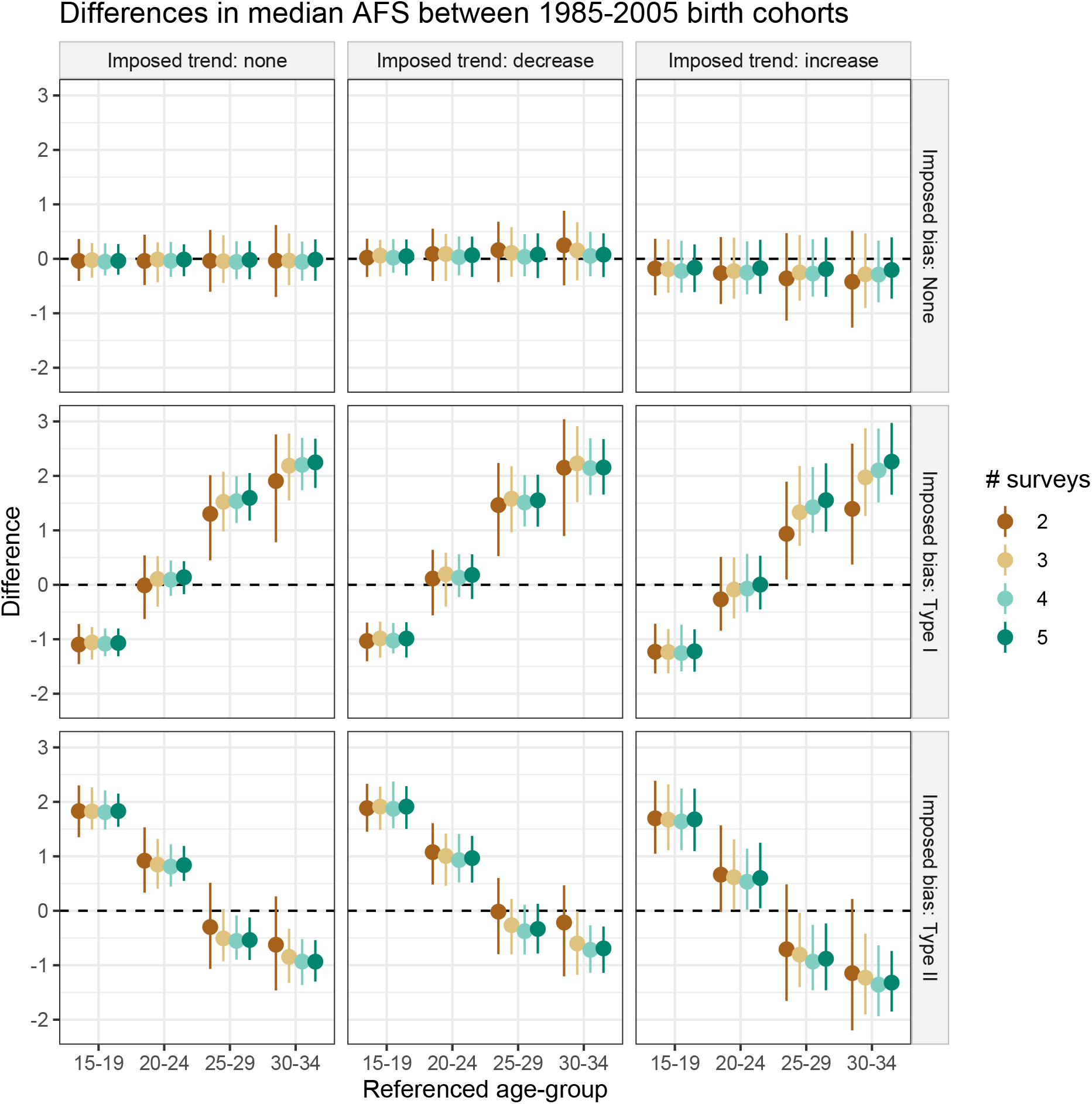
Average difference in the estimated median AFS in the 2005 birth cohort. Different scenarios of the AFS trend, number of surveys, type of biases (none or a logistic function of age). The age’s effect reference group illustrates the effect on the difference when the corresponding average age coefficients of age-group was used in generating the AFS distribution.

